# ASSOCIATIONS BETWEEN ADVERSE CHILDHOOD EXPERIENCES AND INTIMATE PARTNER VIOLENCE WITH POSTPARTUM DEPRESSION AMONG WOMEN IN CENTRAL REGION OF VIETNAM: A PROSPECTIVE COHORT STUDY

**DOI:** 10.1101/2025.06.25.25330312

**Authors:** Trung Tran Dinh, Binh Thang Tran, Thang Vo Van, Vu Huy Nguyen Quoc, Duong Le Dinh, Tao Tran Thi, Kathy Trang, Kathleen M. Baggett, Kathryn M. Yount

## Abstract

**Purpose:** This study used longitudinal data to explore the pathways linking ACEs, IPV during pregnancy (p-IPV), and postpartum IPV (IPV-12) with PPD at 6 and 12 months postpartum in central Vietnam.

**Methods:** A prospective cohort study was conducted, recruiting 942 women in their last trimester of pregnancy in Da Nang, Vietnam, and following them up to 12 months postpartum. ACEs were assessed using the 10-item Adverse Childhood Experiences Questionnaire (ACE-Q) at baseline. IPV (psychological, physical, sexual) was measured using the Revised Conflict Tactics Scales (CTS2) during pregnancy (p-IPV) and at 12 months postpartum (IPV-12). PPD was assessed using the Patient Health Questionnaire-9 (PHQ-9) at 6 months (PPD-6) and 12 months (PPD-12) postpartum. Mediation analysis using the Karlson-Holm-Breen (KHB) method was performed to examine direct and indirect effects.

**Results:** Loss to follow-up was 20.0% at 6 months (n=753) and 15.3% at 12 months (n=638). Mediation analysis indicated that ACEs influenced PPD-12 (Total Effect = 0.536) and that p-IPV, PPD-6, and IPV-12 fully mediated this relationship, accounting for 85.44% of the total effect. IPV-12 emerged as the strongest mediator (71.74%), followed by PPD-6 (7.77%) and p-IPV (5.93%). Specific forms of IPV (psychological, physical, sexual) also demonstrated significant mediating roles in various pathways.

**Conclusion:** This study highlights the critical role of exposure to IPV in the relationship between ACEs and PPD. Recent exposure to IPV (at 12 months postpartum) may act as a particularly potent mediator in this relationship. Community-based screening for ACEs and IPV is recommended as an integral component of PPD prevention strategies.

## INTRODUCTION

Postpartum depression (PPD) refers to the onset of depressive episodes among postpartum women and is recognized as the most common mental health disorder (Amer et al., 2024). An estimated 17.2% of the world’s mothers have experienced PPD (Wang et al., 2021). Notably, the prevalence rate of PPD varies by income and geographic development, which a significantly higher rate being observed in lower-income countries (Wang et al., 2021). From 2020 to 2021, during the COVID-19 pandemic, Asian/Pacific Islanders exhibited the largest relative increase in the prevalence of PPD, from 3.6% to 13.8% or a 280% increase (95% CI: 220-350%) (Getahun et al., 2023). Growing evidence indicates that PPD adversely affects the mother, the infant, and their family relationship. Depressed mothers seem to exhibit more negative emotions, little verbal communication, and less use of preventive services for their infants. Compared to the infants of non-depressed mothers, those of depressed mothers have exhibited impaired maternal–child interactions, poorer physical growth, more behavioral problems, and lower cognitive development (Klainin & Arthur, 2009). Thus, PPD represents a notable concern given its high prevalence and consequences globally.

Experiences of adversity in childhood have an important influence on mental health in adulthood. Adverse Childhood Experiences (ACEs) refer to cumulative retrospective self-reports of childhood adversity including abuse, neglect, and/or family dysfunction before the age of 18, and their accumulation increases the risk of depression in postpartum women (Racine et al., 2021). A study with 16,831 female participants in Iceland demonstrated a positive association between total number of ACEs and perinatal depression (PR 1.11 per ACE, 95% CI: 1.10–1.11, even in women did not have psychiatric comorbidities (PR 1.13, 95% CI: 1.11–1.14) (Bränn et al., 2023). Furthermore, maternal ACEs was shown to indirectly affect child development through maternal stressful events during pregnancy, as well as pre- and postnatal mental health challenges in another prior study in Taiwan (Chang, Feng, Chang, Chang, & Lee, 2021). A separate meta-analysis of 15 studies confirmed the influence of ACEs on maternal mental health (Racine et al., 2021).

Intimate partner violence (IPV) is another global health problem and refers to physical, sexual, and/or emotional harm by a current or former partner (Zhu et al., 2024). Exposure to ACEs is a risk factor for IPV (Zhu et al., 2024), and pregnant women with a history of ACEs and/or IPV may be particularly vulnerable to poor health outcomes. An estimated one-third of pregnant women have a history of childhood abuse or neglect, 25% have experienced IPV at some point in their lives, and 4–8% encounter IPV during pregnancy (B. L. Goldstein, Briggs-Gowan, & Grasso, 2021). ACEs and IPV are identified predictors of maternal mental health problems (B. L. Goldstein et al., 2021). Specifically, four in ten women reported experiencing ACEs, while two in ten reported IPV during the index pregnancy, both of which were significantly associated with symptoms of PPD in another cross-sectional study in Dar-es -Salaam (Mahenge, Stöckl, Mizinduko, Mazalale, & Jahn, 2018). Additionally, a previous cross-sectional study in Albania demonstrated that maternal ACEs was positively associated with stress levels and depressive symptoms. Notably, IPV partially and fully mediated the association of ACEs with maternal stress and with depressive symptoms, which highlights a pathway of cumulative effects on maternal mental health resulting from a series of interpersonal violence victimizations across the lifespan (Ramaj & Eisner, 2025).

To our knowledge, the influence of ACEs and IPV on PPD has been examined primarily using cross-sectional designs. Additionally, the direct and indirect pathways at different time points within these relationships remain unclear, and the exact mechanisms are understudied. This research gap raises the question of whether IPV mediates the relationship of ACEs and PPD and whether the timing of IPV exposure is important in the mediation process. To address this gap, our study employs a cohort design in Vietnam, where ACES (Le, Dang, & Weiss, 2022), IPV (UNFPA and the Government of Viet Nam, 2019) and PPD (Nguyen, Hoang, Do, Schiffer, & Nguyen, 2021)(Van Vo, Hoa, & Hoang, 2017) are all reported to be prevalent. The study is guided by two primary research questions.

### Research questions

1. *What is the total effect of Adverse Childhood Experiences (ACEs) on Postpartum Depression (PPD) at 6 and 12 months postpartum among pregnant women in Vietnam?*
2. *To what extent does the total effect of ACEs on PPD operate through women’s exposure to IPV during and after pregnancy?*

## METHODS

### Study design

A prospective cohort study.

### Participants

The study subjects were pregnant women recruited at the end of pregnancy and followed up continuously until their children reached 12 months of age. Women were eligible to participate if they had the following characteristics: in the last 3 months of pregnancy, with a gestational age of 29 to 40 week; 18 and 49 years having a permanent residence registration or a long-term temporary residence certificate (minimum 12 months) in Da Nang city. We excluded those who have a plan of moving out of Da Nang City within the next 12 months of follow-up; and unable to communicate effectively or provide reliable information for research purposes (due to severe language barrier, severe cognitive impairment).

### Sample size

In this project, we utilized the data from a large-scale perspective cohort study. Of the 1235 women invited, 986 were interviewed at their third trimester of pregnancy, resulting in a baseline participation rate of 80%. By then, after medical record review, a total of 942 pregnant women were eligible to include at the baseline analysis. Participants were then followed at 6 and 12 months postpartum, from October 2022 to December 2023.

### Variables and Measurements

#### Adverse childhood experiences (ACEs)

ACEs are negative experiences during childhood that a person may go through, including events such as family violence, sexual abuse, lack of care, abandonment, or witnessing heart-wrenching events like divorce or a family member’s incarceration. We use the Adverse Childhood Experiences-Questionnaire (ACE-Q) 10-item scale (5 family dysfunction items and five exposures to violence items) to assess experiences before the age of 18 in pregnant women. (Felitti et al., 1998; Zarse et al., 2019). ACE-Q has been widely used in various regions and with different target groups since 1998. To date, the instrument has been translated and utilized in Vietnam (Hoang et al., 2024).

In the ACE-Q, participants self-reported whether (yes=1) or not (no=0) they had experienced each adversity. The total score ranges from 0 to 10. Pregnant women who have any experiences from the questionnaire was considered to have adverse childhood experiences (Hoang et al., 2024). In this study, we measured ACE-Q once during the last 3 months of pregnancy (baseline). In our preliminary analysis of data at baseline (T0, n=85), Cronbach’s alpha of 10-items of ACE-Q was 0.89.

#### Intimate Partner Violence (IPV)

IPV was defined as exposure to sexual, physical, or psychological violence by current spouse. We used the Revised Conflict Tactics Scales (CTS2) to assess IPV and its forms. Basically, CTS2 comprised 21 questions measures of individual engagement in or experience of physical or psychological violence with an intimate partner. Each question was rated in different levels from one to five, corresponding to: 1: 1 time, 2: 2-5 times, 3: times; 4: times, and more than 5 times. And a score of 0 is assigned if the subject does not exhibit any acts of violence during pregnancy with two levels being “never” and “not during pregnancy but before” (Vy, 2019).

#### Psychological violence (PsV)

PsV was measured using four yes-no items: “Has your partner/spouse ever: 1) Insulted/Humiliated/Verbally abused; 2) Damaged your reputation/status in front of others; 3) Threatened/Intimidated; 4) Threatened to harm or harm you or someone you love.” If all 4 questions were answered “Never,” it is considered as no psychological abuse; otherwise, it will be classified into the group with psychological abuse. *Physical violence (PhV):* PhV was measured using 11 different items: “Has your partner/spouse ever: 1) Intentionally pushed you; 2) Pushed something onto you; 3) Slapped your face; 4) Thrown something at you; 5) Used an object or their hands to hit you; 6) Kicked you; 7) Scratched or pulled you; 8) Hit you with a fist; 9) Choked you; 10) Caused burns; 11) Used a knife or gun to threaten or assault.” If all 11 questions were answered “Never,” it is considered as no physical abuse; otherwise, it will be classified into the group with physical abuse (PA).

#### Sexual violence (SV)

SV was measured using 6 different items: “Has your partner/spouse ever: 1) Demanded sexual intercourse from you; 2) Forced sexual intercourse with you; 3) Used force to engage in sexual intercourse; 4) Forced oral sex; 5) Forced anal sex; 6) Used objects for sexual intercourse.” If all 6 questions were answered “Never,” it is considered as no sexual abuse; otherwise, it will be classified into the group with sexual abuse.

The CTS2 scale shows good reliability (Cronbach’s alpha = 0.81 and McDonald’s omega=0.80, and ICC=0.91 in our previous study (Trần Đình Trung, Nguyễn Vũ Quốc Huy, & Võ Văn Thắng, 2024). Furthermore, reliability coefficients for its subtypes were reported as: Psychological Abuse = 0.83, Physical Abuse = 0.76, and Sexual Abuse = 0.78 (Trần Thị Nhật Vy, 2019).

In this longitudinal study, we measured IPV using CTS2 at two points: the first was at the last 3 months of pregnancy (abbreviation: p-IPV), and the second was at 12 months after birth (abbreviation: IPV). p-IPV was IPV asked with respect to the entire period of pregnancy at the point of their third trimester of pregnancy. Questions for IPV-12 pertained to the 12 months subsequent to their child’s birth.

#### Postpartum depression (PPD)

PPD indicates psychological and emotional symptoms that women may experience after giving birth. We use the Patient Depression Questionnaire (PHQ9) scale to assess the level of postpartum depression in mothers. These questions focus on symptoms such as loss of interest, feelings of sadness, sleep problems, fatigue, self-blame, difficulty concentrating, lack of confidence, loss of pleasure, and suicidal thoughts. A previous study in Vietnam indicated as good reliability (Cronbach alpha reliability of 0.79) (Do et al., 2021).

The PHQ-9 consists of 9 items, with responses to each item rated on a Likert scale from 0 to 3 to capture the frequency of the symptoms in the prior 2 weeks/months: corresponding to: 0 “Not at all”, 1 “Several days”, 2 “More than half the days”, and 3 “Nearly every day.”. The item responses are summed to create a total possible score ranging from 0 to 27.

The total score on the PHQ-9 can be interpreted as follows: 0-4: no depression symptoms; 5-9: Mild depression; 10-14: Moderate depression; 15-19: Moderately severe depression; and 20-27: Severe depression (Kroenke, Spitzer, & Williams, 2001). In this study, we administered the PHQ-9 at two points: 6 months after birth and 12 months after birth.

### Data collection

The data collection was conducted in two phases (Figure 1). The initial phase, a cross-sectional data collection from October 2022 to April 2023, focused on recruiting participants. This involved securing necessary permissions from local health authorities and closely collaborating with commune health centers. Potential participants were identified and screened based on predefined criteria using health center records. Eligible women then were invited for private interviews at the health centers, where informed consent was obtained before administering standardized questionnaires to gather baseline data.

**Figure 1.**
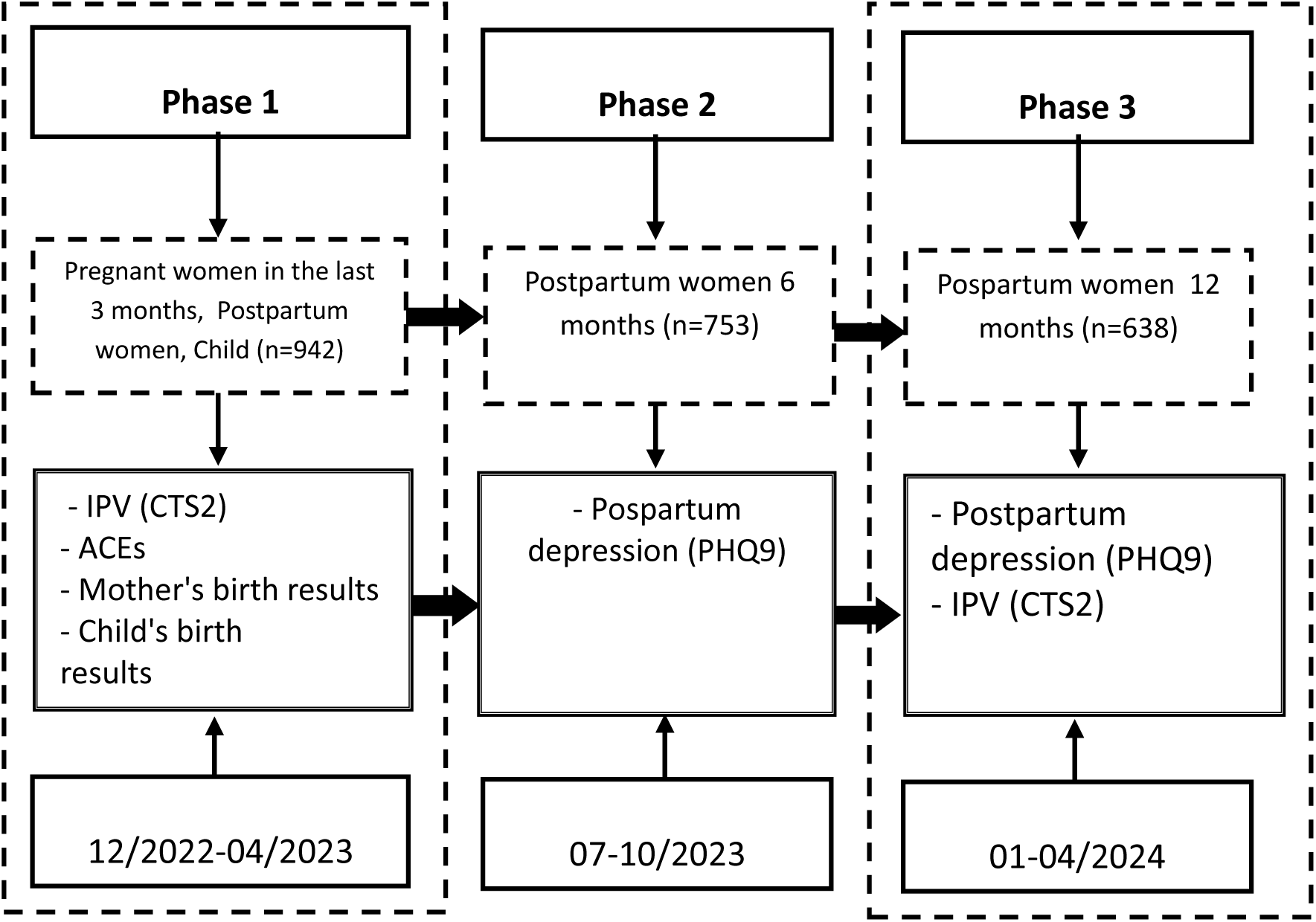
Conceptual Framework.

The second phase involved a longitudinal follow-up from January 2023 to April 2024 of the participants recruited in the first phase. The initial postpartum follow-up gathered data on maternal and infant birth outcomes. This was primarily done by reviewing hospital records, supplemented by records from health centers and city health departments, with data collected immediately at birth. Follow-ups at 6 and 12 months postpartum involved contacting participants for in-person interviews at health centers. During these interviews, standardized follow-up questionnaires were used to assess maternal mental health and child development, with interviewers receiving specialized training for administering and scoring the child development assessment tool. Throughout both phases, rigorous supervision was maintained to ensure the integrity, accuracy, and confidentiality of the collected data.

### Statistical analysis

Descriptive statistics were performed to present the overall characteristics of the cohort, using means and standard deviations (SD) for continuous data as well as observed counts with percentages for categorical data. To define the relationship between ACEs, IPV, and PPD, mediation analysis was performed using the Karlson-Holm-Breen (KHB) method (Joreskog & Sorbom, 1993; Kohler, Karlson, & Holm, 2011), which is appropriate for nonlinear mediation models (Figure 2). The relationships between ACEs, IPV and PPD were assessed by estimating the total effect, direct effects and the indirect effects for each hypothesis, as shown in Figure 1. Figure 2 the path analysis to test the direction of association ACE and PPD at 12 months, with both prenatal IPV (p-IPV) and IPV at 12 months (IPV-12) and PPD at 6 month or their subtypes acting as mediators. Statistical significance was set at α = 0.05. Data analysis was utilized Stata version 16 software.

**Figure 2.**
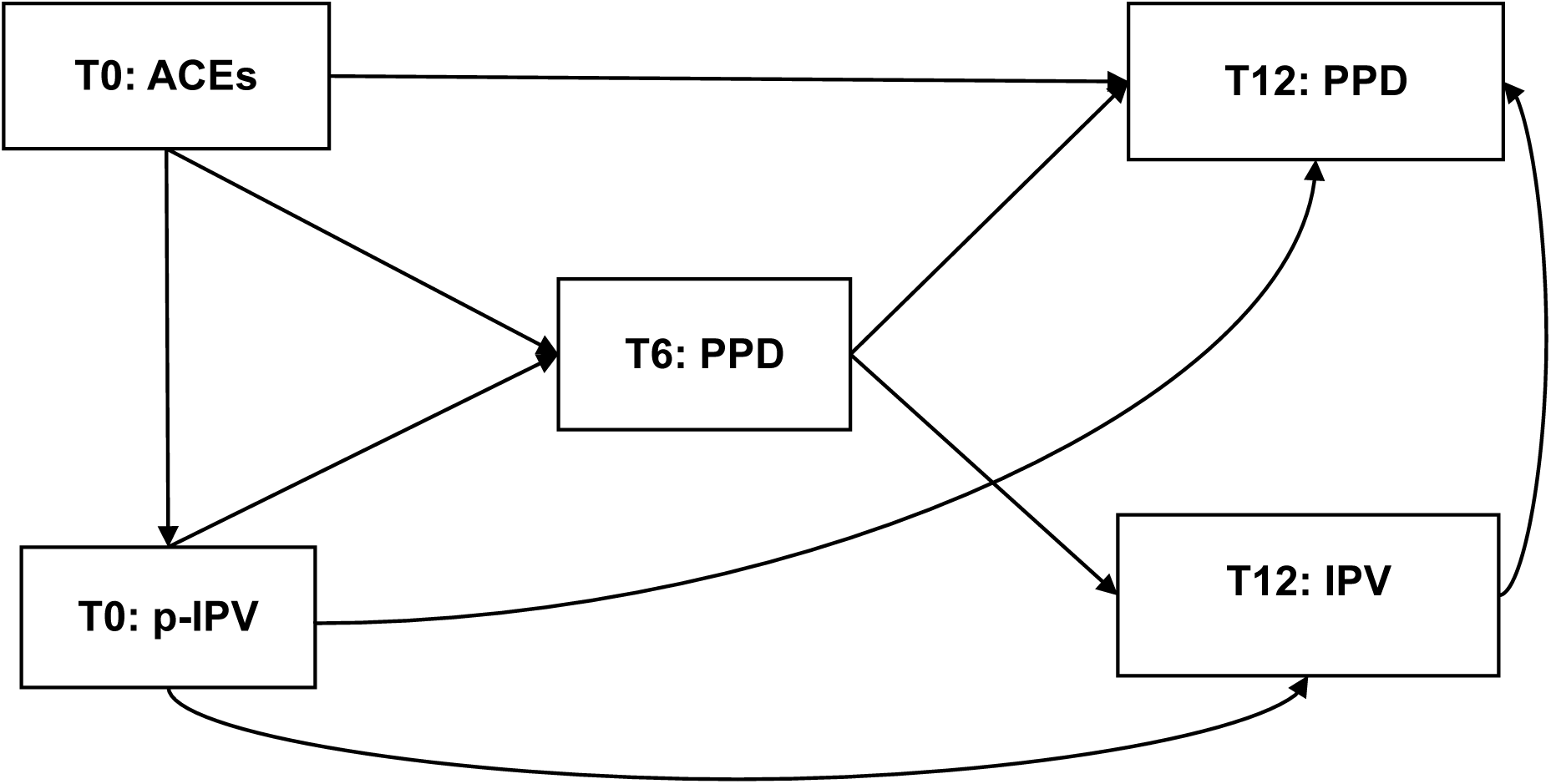
Path model assessing the relationship between ACE, IPV and PPD during the period. IPV: Intimate Partner Violence; p-IPV: Intimate Partner Violence During PregnancyACEs: Adverse Childhood Experiences; PPD: Postpartum Depression; T0: baseline during last 3 months of pregnancy duration; T6: at 6 months postpartum; T12: at 12 months postpartum.

### Ethics consideration

This study received ethical approval from the Biomedical Research Ethics Committee of Hue University of Medicine and Pharmacy (H2020/503). And Biomedical Research Ethics Committee of Da Nang University of Medical and Pharmacy (DUMTP-2023-126). Throughout the research, strict adherence to WHO guidelines on ethical and safety considerations for domestic violence research was maintained.

## RESULTS

### Participant Characteristics

In the initial phase, 942 eligible participants were recruited (participation rate: 76%). At the 6-month postpartum follow-up, 753 mothers were retained, indicating a 20.0% attrition rate from baseline. For the 12-month follow-up, 638 mothers were assessed, showing a 15.3% attrition rate since the first follow-up. The final number of participants retained for the 12-month assessment was 638, resulting in an overall participation rate of 57.7%.

Table 1 presents the baseline and follow-up demographic data for the study participants. Initially, 942 mother-child dyads were enrolled (T0). At baseline, the average maternal age was 30.3±5.3 years, with 70.6% (665) of mothers aged between 25-35 years. A majority of mothers (56.6%, 533) had completed high school and above levels of education. Husbands’ mean age was 32.7±5.3 years, and 57.2% (539) had attained upper high education. The children comprised 53.7% (506) males and 46.3% (436) females. By the 6-month follow-up (T6), 753 participants remained, and 638 participants were present at the 12-month follow-up (T12). The 304 participants lost to follow-up by T12 had a mean baseline maternal age of 30.0±5.4 years, and 53.6% had high school and above education, broadly similar to the overall baseline cohort.

**Table 1.** Baseline and followed-up data on the general characteristics of participants.

|  | **T0**<br>**N=942** | **T6**<br>**N=753** | **T12**<br>**N=638** | **T6 vs T0**<br>**Missing**<br>**rate**<br>**n=189** | **T12 vs T6**<br>**n=115** | **T12 vs T0**<br>**n=304** |
| --- | --- | --- | --- | --- | --- | --- |
| **Mother's age** | $30.3 \pm 5.3$ | $30.1 \pm 5.2$ | $30.4 \pm 5.2$ | $30.7 \pm 5.4$ | $28.7 \pm 5.2$ | $30.0 \pm 5.4$ |
| 18-<25 | 131 (13.9) | 108 (14.3) | 79 (12.4) | 23 (12.2) | 29 (25.2) | 52 (17.1) |
| 25-35 | 665 (70.6) | 533 (70.8) | 458 (71.8) | 132 (69.8) | 75 (65.2) | 207 (68.1) |
| > 35 | 146 (15.5) | 112 (14.9) | 101 (15.8) | 34 (18.0) | 11 (9.6) | 45 (14.8) |
| **Mother's**<br>**education** |  |  |  |  |  |  |
| Less than high<br>school | 222 (23.6) | 173 (23.0) | 141 (22.1) | 49 (25.9) | 32 (27.8) | 81 (26.6) |
| High school | 187 (19.9) | 150 (19.9) | 127 (19.9) | 37 (19.6) | 23 (20.0) | 60 (19.7) |
| More than<br>high school | 533 (56.6) | 430 (57.1) | 370 (58.0) | 103 (54.5) | 60 (52.2) | 163 (53.6) |
| **Husband's age** | $32.7 \pm 5.3$ | $32.5 \pm 5.3$ | $32.8 \pm 5.3$ | $33.2 \pm 5.3$ | $31.1 \pm 5.3$ | $32.4 \pm 5.4$ |
| <25 | 79 (8.4) | 67 (8.9) | 54 (8.5) | 12 (6.3) | 13 (11.3) | 25 (8.2) |
| 25-35 | 619 (65.7) | 495 (65.7) | 417 (65.4) | 124 (65.6) | 78 (67.8) | 202 (66.4) |
| > 35 | 244 (25.9) | 191 (25.4) | 167 (26.2) | 53 (28.0) | 24 (20.9) | 77 (25.3) |
| **Husband's**<br>**education** |  |  |  |  |  |  |
| Less than high school | 70 (7.4) | 56 (7.4) | 44 (6.9) | 14 (7.4) | 12 (10.4) | 26 (8.6) |
| High school | 333 (35.4) | 274 (36.4) | 238 (37.3) | 59 (31.2) | 36 (31.3) | 95 (31.3) |
| More than high school | 539 (57.2) | 423 (56.2) | 356 (55.8) | 116 (61.4) | 67 (58.3) | 183 (60.2) |
| **Child sex** |  |  |  |  |  |  |
| Males | 506 (53.7) | 410 (54.4) | 348 (54.5) | 96 (50.8) | 62 (53.9) | 158 (52.0) |
| Females | 436 (46.3) | 343 (45.6) | 290 (45.5) | 93 (49.2) | 53 (46.1) | 146 (48.0) |

### Prevalence Rates of Exposure to ACEs and IPV

Table 2 details the prevalence of ACEs, IPV, and PPD across the study waves. ACEs data, collected at baseline (T0, N=942), showed that 38.4% (362) of mothers reported no ACEs, while 13.5% (127) reported two ACEs, and a cumulative 32.3% reported experiencing five or more ACEs. IPV during pregnancy (p-IPV at T0) was reported by 31.2% (294) of women. At 12 months postpartum (IPV-12, n=638), IPV was reported by 29.8% (190) of women. Psychological violence was the most frequently reported IPV subtype at both T0 (25.1%, 236) and T12 (24.6%, 157). Mean PPD scores were 1.37±1.45 at T0, 3.76±4.69 at T6, and 3.30±4.20 at T12. Clinically significant PPD was nil at T0 but affected 30.7% (196) of mothers at T6 and 25.4% (191) at T12

**Table 2:**
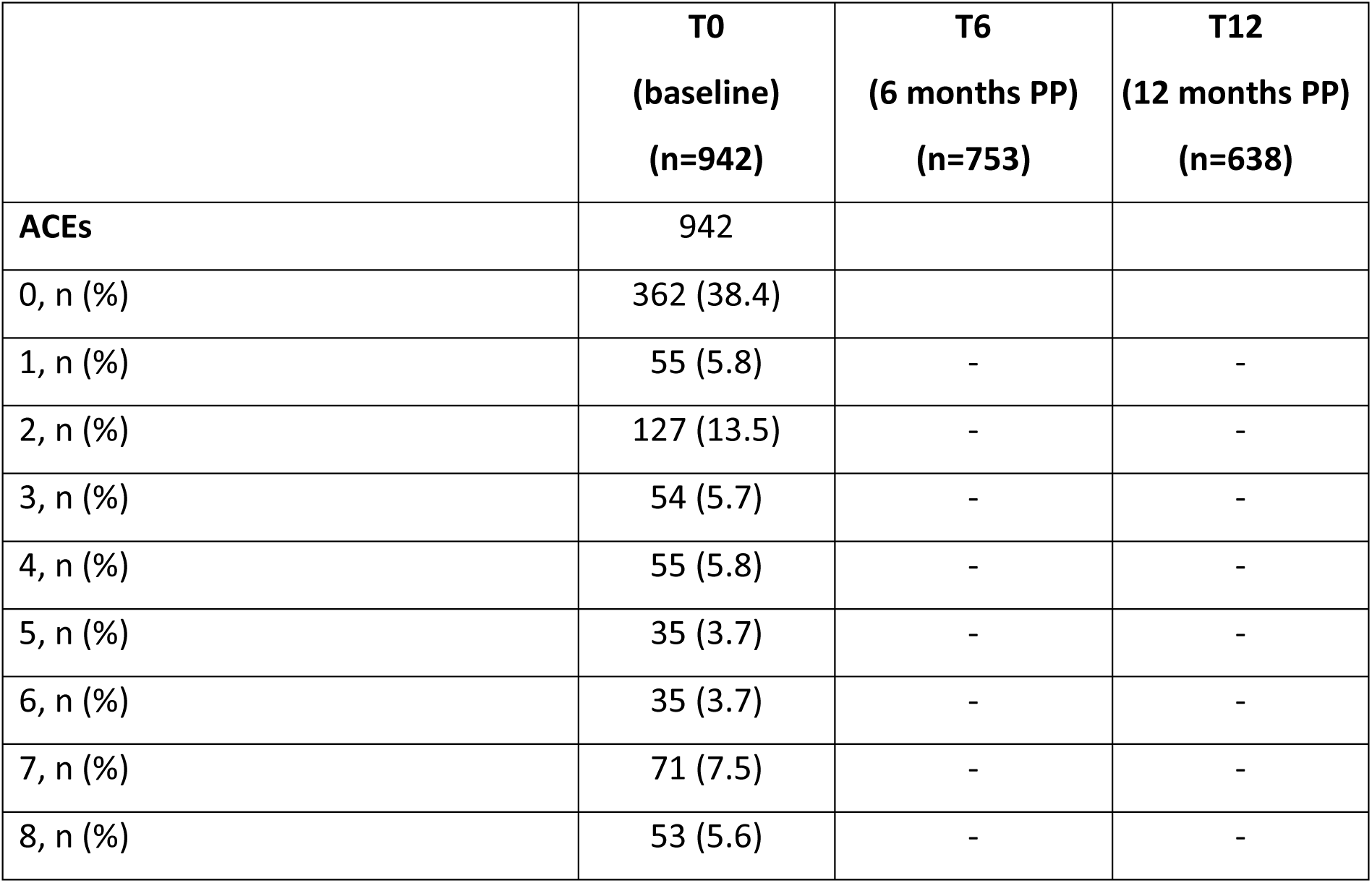
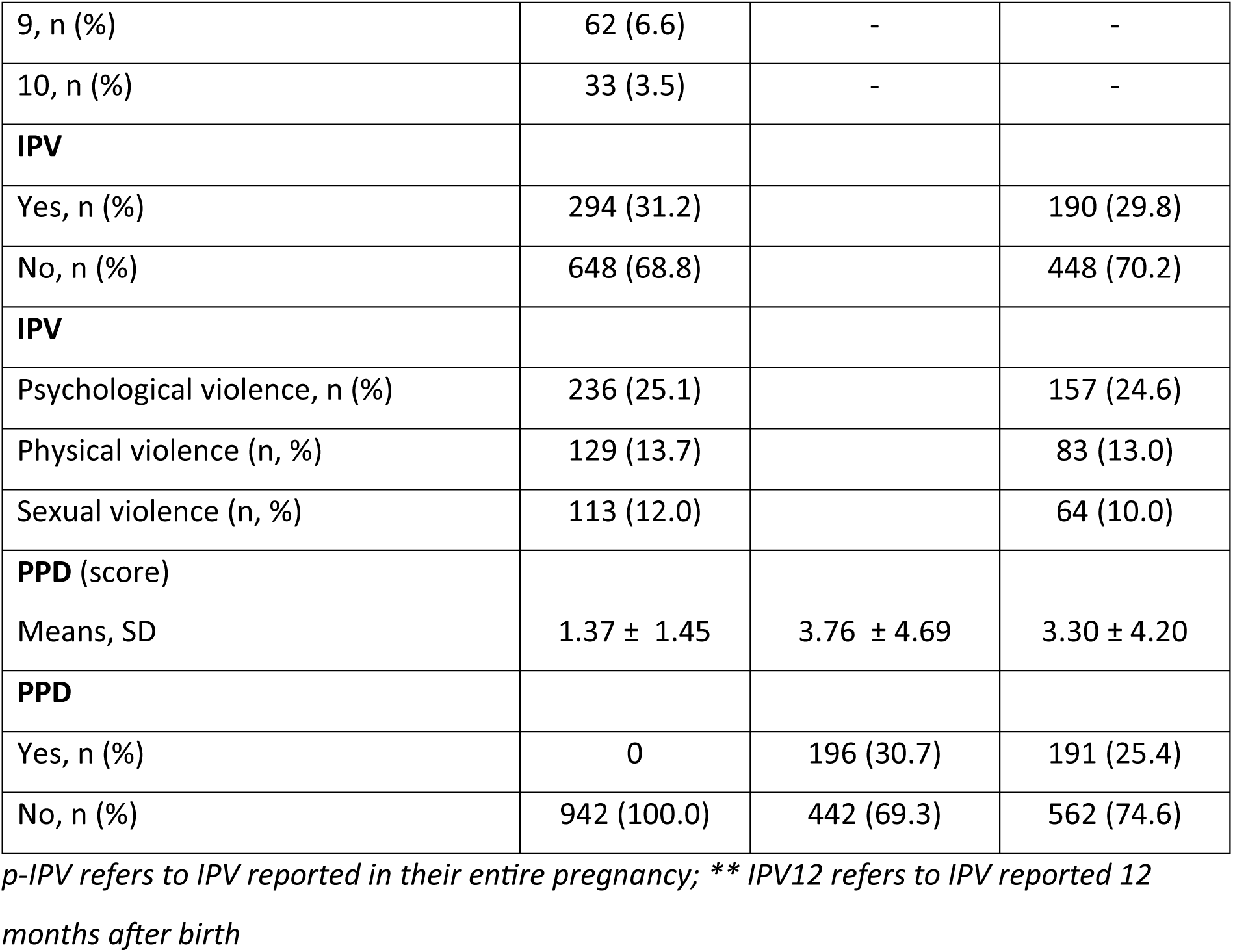
ACEs, IPV, subtype IPV, PPD at waves of study.

### Mediation Analysis

The total effect of ACEs on PPD-6, 0.555 (95% CI = 0.421, 0.689), could be decomposed into a direct effect, 0.376 (95% CI = 0.255, 0.496), and an indirect effect, 0.179 (95% CI = 0.055, 0.304). In relative terms, the indirect effect constituted 32.3% of the total effect.

The total and direct effects of the pattern of p-IPV on the PPD-6 were 0.527 (95% CI = 0.404, 0.650) and 0.271 (95% CI = 0.163, 0.379), and the indirect effect of IPV forms was 0.256 (95% CI = 0.103, 0.409), with 48.57% of the total effect being mediated. Breaking down the indirect effect into its components, a substantial part of the indirect effect was via psychological IPV (22.84%), followed by physical IPV (13.86%) and then sexual IPV (11.88%).

In general, across all models, partial mediation by p-IPV and IPV-12 was observed (Table 3). Model 6 demonstrated complete mediation, with its indirect effect on PPD-12 being significant and accounting for 85.44% of the mediation

**Table 3.**
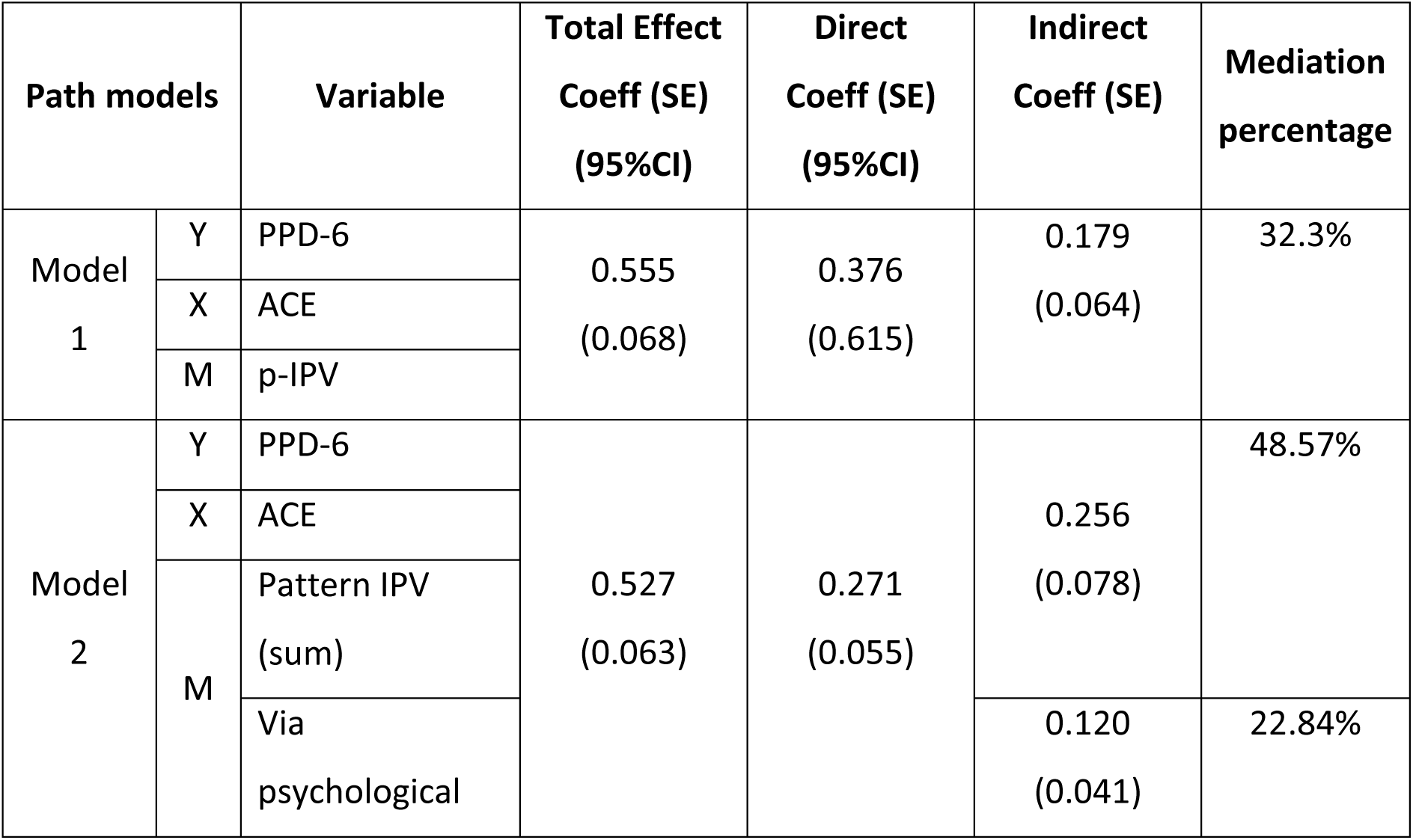
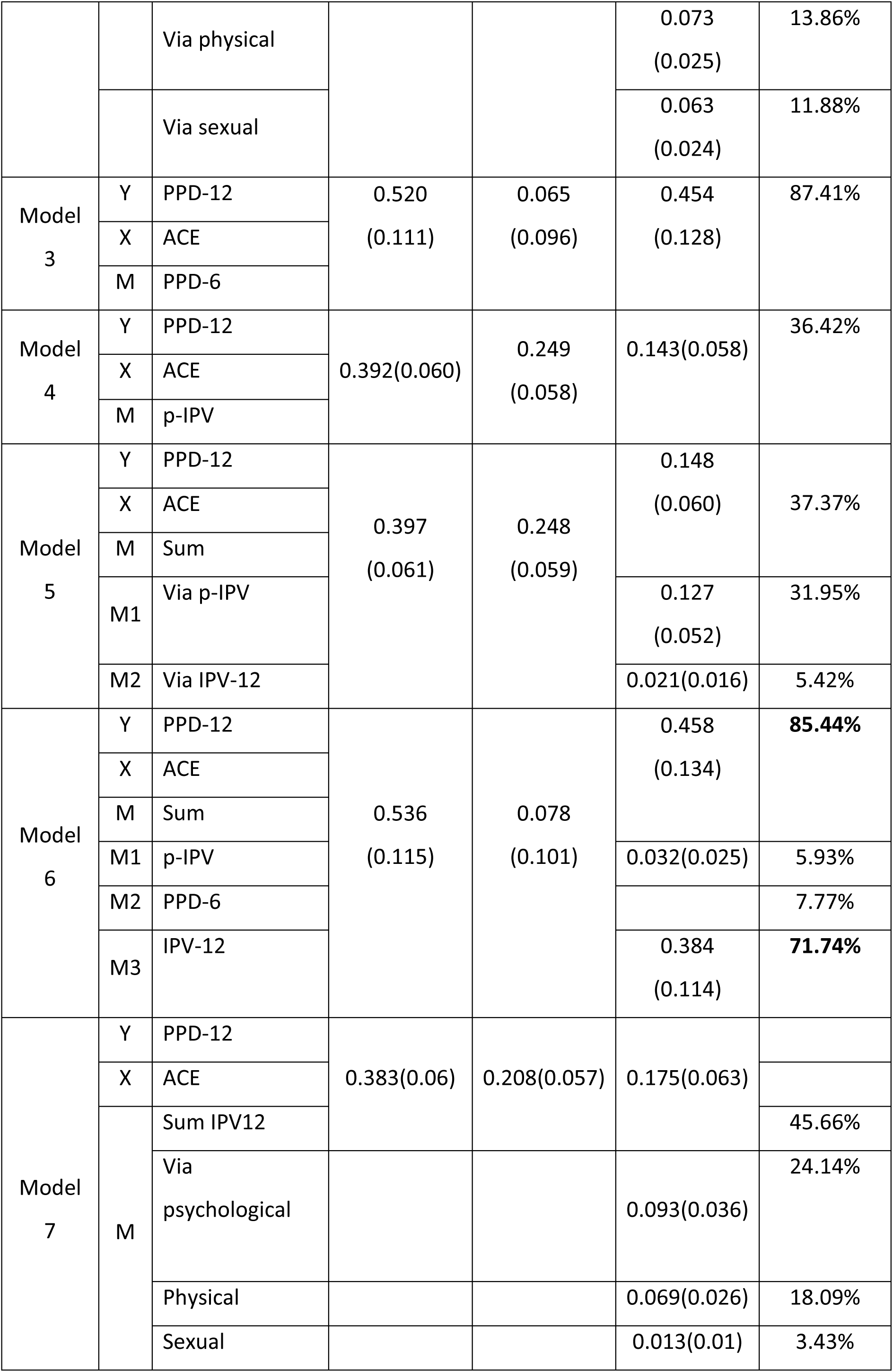
Mediation analysis of factors associated with PPD using the Karlson-Holm-Breen (KHB) method, 638 pregnant women followed for 12 months post-partum, clinics in Da Nang city, Vietnam.

## DISCUSSION

### Summary of Findings and Interpretation

In our cohort study of 942 women, ACEs had a significant total effect on postpartum depression at 6 months, with 32.3% of this effect occurring indirectly through mediators. Similarly, IPV also significantly impacted PPD6, with 48.57% of the total effect mediated. Most effects showed partial mediation, except the influence of ACEs on PPD at 12 months with complete mediation through different pathways. Specifically, the total effect of ACE on PPD at 12 months was 0.536, with full mediation by p-IPV, PPD-6, and IPV-12, accounting for 85.44% of the total effect. The strongest mediation was observed for IPV-12 (71.74%), followed by PPD-6 (7.77%) and p-IPV (5.93%).

Our findings corroborate and add to previously published findings regarding these relationships. A previous retrospective cohort of 735 low-income women in Wisconsin demonstrated the partial indirect effect of ACEs on PPD, mediated by IPV, perceived stress, and antenatal depression (Mersky & Janczewski, 2018). Similarly, in a previous cross-sectional study in Tirana, Albania. ACEs had a positive indirect influence on depressive symptoms that was entirely mediated by positive pathways from ACEs to IPV and IPV to depressive symptoms (Ramaj & Eisner, 2025). The indirect effect of ACEs on postpartum depression was also confirmed in a study using data of South Dakota Pregnancy Risk Assessment Monitoring System, which included 2,343 postpartum women (E. Goldstein & Brown, 2023). However, in prior studies, the mediating role of IPV at specific time points—including p-IPV and IPV-12-has not been fully explored in the ACE–PPD relationship. Our study highlights the full mediating effects of p-IPV, and especially IPV-12 in the link between ACEs and PPD at 12 months postpartum.

These results imply that exposure to ACEs increases the risk of exposure to intimate partner violence in adulthood, and in turn, an increased risk of poor mental health (Mersky & Janczewski, 2018). Specifically, in comparison with participants without ACEs, those with at least one ACE experience have had a 215.5% higher risk of IPV in other studies (Tian, Zhang, Li, Wu, & Wang, 2024). Possible explanations may be proposed for that. First, an individual’s experiences may influence perception, attitudes, behaviours and expectations in intimate relationships in adulthood, thus, individuals may unconsciously repeat or mimic childhood patterns in partnerships. ACE may impact on perceptions, leading to greater acceptance of violence and the misperception of violence as a legitimate form of discipline or at least something to be tolerated. Children are more vulnerable because they learn behavior by observing violence at home, thus, they are more likely to accept violence and imitate these violent behaviors in future intimate relationships(Tian et al., 2024). Similarly, violence acceptance is common among Vietnamese women, with IPV often perceived as a normal part of marital life - particularly when it is infrequent or less severe. Women are more likely to seek help only when IPV becomes frequent or severe (James-Hawkins, Hennink, Bangcaya, & Yount, 2021).

Importantly, our findings highlight the influence of ACEs on IPV, which in turn affects PPD, aligning with existing evidence. A significant positive trend was observed in mental health risks as the number of ACEs increased. Pregnant women with 1-2 ACEs had a 2.4 to 3.2 times higher risk of experiencing anxiety disorders, depressive disorders, or p-IPV. Those with 3 or more ACEs faced a 3.1 to 4.7 times higher risk of anxiety disorders, depressive disorders, depressive symptoms, or p-IPV (Young-Wolff et al., 2019). The physiological stress response system through epigenetic mechanisms and is linked to changes in the volume of stress-sensitive brain regions might be changed by prolonged exposure to stress, especially during the neurodevelopmentally sensitive period from infancy through early childhood. ACE was demonstrated to have an association with molecular markers of disease risk and aging such as an increase in mitochondrial DNA copy number, which may affect women during the prenatal period, which is especially significant given the potential for intergenerational transmission of ACEs through these biological pathways (Young-Wolff et al., 2019).

Notably, current evidence documented an association of any IPV exposure and sign development of postpartum depression (Ankerstjerne et al., 2022). The link between IPV to PPD can be explained as follows. First, PPD has ben recognized to be associated with several socioeconomic, psychological, and cultural factors. Exposure to IPV can disrupt the balance between individual resources and environmental demands, reducing resilience, increasing vulnerability to mental health issues, and ultimately contributing to the onset of depression (Ankerstjerne et al., 2022). Additionally, IPV can undermine a victim’s trust in others, increase fear, influence coping styles, and lead to greater social isolation—all of which may heighten the risk of depression (Ankerstjerne et al., 2022). Notably, we consider IPV over an extended period—from during pregnancy to several months postpartum—offering a comprehensive view of its mediating role in the relationship between ACEs and PPD. There was little or no awareness on this potential mediation in existing literature. IPV at 12 months postpartum emerged as the strongest mediator for the association of ACEs with PPD at 12 months in our study. This finding aligns with previous research highlighting recent IPV exposure as a strong independent predictor of PPD within the first year after childbirth. These results highlight the importance of the timing of IPV exposure and the need to strengthen screening and referral efforts for IPV to reduce postpartum mental health morbidity (Valentine, Rodriguez, Lapeyrouse, & Zhang, 2011).

### Limitations and Strengths of Study

This study possesses notable strengths, primarily its prospective cohort design, which is one of the few to longitudinally examine the complex pathways linking ACEs, IPV at specific time points (during pregnancy and 12 months postpartum), and PPD up to 12 months postpartum in central Vietnam. This approach allowed for a nuanced exploration of the mediating role of IPV, particularly highlighting that IPV experienced 12 months postpartum (IPV-12) was the most potent mediator in the relationship between ACEs and PPD at 12 months. The use of validated instruments like the ACE-Q, CTS2, and PHQ-9, with reported reliability in the Vietnamese context or in preliminary analyses, also adds to the study’s rigor.

However, a significant limitation is the substantial loss to follow-up. The study experienced a 20.0% attrition rate at the 6-month follow-up and a further 15.3% attrition by the 12-month follow-up, resulting in an overall participation rate of 57.7% at the final assessment point. This high attrition was attributed to local customs, social norms discouraging postpartum visits, and the transient nature of young families. Such attrition could potentially introduce bias and affect the generalizability of the findings, as the characteristics of those lost to follow-up might differ from those who remained in the study.

### Recommendations for Future Research and Policy

The profound impact of ACEs and IPV on maternal PPD in central Vietnam necessitates urgent action. Research reveals a clear longitudinal pathway where ACEs increase vulnerability to IPV, both during pregnancy (p-IPV) and postpartum (IPV-12), which in turn significantly elevates PPD risk. Notably, IPV at 12 months postpartum (IPV-12) is a potent mediator, accounting for a large portion of ACEs’ effect on PPD. Given the high prevalence of these adversities—with many women reporting ACEs, experiencing IPV, and suffering from PPD —a comprehensive strategy combining targeted research, robust policy, and strengthened practice is crucial to interrupt this cascade and improve maternal and child well-being.

Future research must prioritize several key areas. *First*, a deeper understanding of the etiological pathways within Vietnam’s cultural context is needed, including exploring cultural influences, long-term intergenerational impacts, and protective resilience factors. *Second*, developing and evaluating culturally adapted, feasible, and acceptable screening strategies for ACEs and IPV within routine maternal care is essential, considering optimal timing and tools. *Third*, innovating and rigorously testing tailored intervention models is critical. This includes IPV prevention and mitigation strategies (such as couple-based interventions where safe, and community or healthcare provider-led initiatives), interventions for sub-threshold and clinical PPD (potentially leveraging scalable approaches like technology-supported mindfulness or stepped-care models), and combined interventions that concurrently address IPV and mental health. *Finally*, implementation science is vital to ensure these effective strategies can be integrated into the existing health system, addressing barriers and training providers.

Translating these research insights into policy and practice requires decisive steps. Key among these is integrating universal, confidential screening for ACEs and IPV into routine Maternal and Child Health (MCH) services, linked to clear referral pathways. Concurrently, access to a spectrum of evidence-based, culturally appropriate interventions must be expanded. This includes comprehensive IPV prevention and support services, a range of maternal mental health services for PPD from prevention to treatment, and integrated care models for co-occurring IPV and PPD. Building workforce capacity through comprehensive training and fostering multi-sectoral collaboration are foundational. Ultimately, addressing broader social determinants, such as challenging harmful gender norms and strengthening legal protections, is imperative to break intergenerational cycles of adversity and promote the long-term health of women and families in Vietnam.

## CONCLUSION

Our study provides evidence that the timing of IPV exposure plays a critical role in the association between ACEs and PPD. This suggest a chain of exposure to violence, where the most recent episode is the outcome of a cascade of prior adversities. Each experience is interconnected, collectively contributing to the development of postpartum depression. These findings highlight influence of cumulative exposure to violence on women’s PPD, emphasizing the need for community-based screening of both ACEs and IPV history as part of depression prevention efforts.

## Acknowledgements

We would like to gratefully acknowledge the support of the CONVERGE project, PI by Prof. Kathryn M. Yount and Prof. Le Minh Giang. And we would like to extend our sincere thanks to Prof. Le Minh Giang and Prof. Hoang Thi Hai Van for their invaluable guidance and support throughout this research.

## Author contributions

Conceptualization: TDT, VVT, NVQH, TBT, KMY, KT, KMB. Methodology: TDT, VVT, TBT, KMY, KT, KMB. Formal analysis: TDT, TBT, LDD. Investigation: TDT, TBT, LDD. Resources: TDT, VVT, NVQH. Data Curation: TDT, TBT, LDD. Writing - Original Draft: TDT, TBT, TTT. Writing - Review & Editing: TDT, TBT, TTT, KMY. Supervision: VVT, NVQH. Funding acquisition: KMY. All authors read and approved the final manuscript.

